# Hippocampal grading provides higher Alzheimer’s Disease prediction accuracy than hippocampal volume

**DOI:** 10.1101/2022.06.29.22275982

**Authors:** Cassandra Morrison, Mahsa Dadar, Neda Shafiee, D. Louis Collins, Alzheimer’s Disease Neuroimaging Initiative

## Abstract

**Background:** Finding an early biomarker of Alzheimer’s disease (AD) is essential to develop and implement early treatments. Much research has focused on using hippocampal volume to measure neurodegeneration in aging and Alzheimer’s disease (AD). However, a new method to measure hippocampal change, known as hippocampal grading, has shown enhanced predictive power in older adults. It is unknown whether this method can capture hippocampal changes at each progressive stage of AD better than hippocampal volume. The goal of this study was to determine if hippocampal grading is more strongly associated with group differences between normal controls (NC), early MCI (eMCI), late (lMCI), and AD than hippocampal volume.

**Methods:** Data from 1666 Alzheimer’s Disease Neuroimaging Initiative older adults with baseline MRI scans were included in the first set of analyses (513 normal controls NC, 269 eMCI, 556 lMCI, and 328 AD). Sub-analyses were also completed using only those that were amyloid positive (N=834; 179 NC, 148 eMCI, 298 lMCI, and 209 AD). We compared seven different classification techniques to classify participants into their correct cohort using 10-fold cross-validation. The following classifiers were applied: support vector machines, decision trees, k-nearest neighbors, error-correcting output codes, binary Gaussian kernel, binary linear, and random forest. These multiple classifiers enable comparison to other research and examination of the most suitable classifier for Scoring by Nonlocal Image Patch Estimator (SNIPE) grading, SNIPE volume, and Freesurfer volume. This model was then validated in the Australian Imaging, Biomarker & Lifestyle Flagship Study of Ageing (AIBL).

**Results:** SNIPE grading provided the highest classification accuracy over SNIPE volume and Freesurfer volume for all classifications in both the full sample and amyloid positive sample. When classifying NC from AD, SNIPE grading provided an accuracy of 89% for the full sample and 87% for the amyloid positive group. Much lower accuracies of 65% and 46% were obtained when using Freesurfer in the full sample and amyloid positive sample, respectively. Similar accuracies were obtained in the AIBL validation cohort for SNIPE grading (NC vs AD: 90% classification accuracy).

**Conclusion:** These findings suggest that SNIPE grading offers increased prediction accuracy compared to both SNIPE volume and Freesurfer volume. SNIPE grading offers promise as a means to classify between people with and without AD. Future research is needed to determine the predictive power of grading at detecting conversion to MCI and AD in amyloid positive cognitively normal older adults (i.e., early in the AD continuum).

**Key points:**

- HC grading may better classify different disease cohorts than HC volume
- Higher prediction accuracy was obtained for HC grading than HC volume
- HC grading offers promise as a method to detect declines in aging and Alzheimer’s

## 1 Introduction

Dementia is a term used to describe a range of disorders that are caused by abnormal brain changes in aging. These abnormal changes impair cognitive functions such as memory, language, and problem-solving that are severe enough to interfere with daily life and independence (Alzheimer’s Association, 2022). The most common form of dementia, Alzheimer’s disease (AD), accounts for 60-80% of dementia cases. AD is a progressive neurodegenerative disorder defined by its underlying pathologies of ß amyloid (Aß42) deposition, pathological tau, and neurodegeneration [AT(N)] (Jack, 2018). Unfortunately, these pathological changes can start years, even decades, before the onset of cognitive symptoms (Sperling et al., 2011). Researchers must thus develop diagnostic tools that can detect AD pathology before too much irreversible neurodegeneration occurs.

Many recent studies have attempted to improve AD classification accuracy using various biomarkers such as cognitive testing, positron emission tomography (PET), cerebrospinal fluid (CSF) assays of Aß42, tau, or magnetic resonance imaging (MRI) changes. For example, using episodic memory test such as the California Verbal Learning Test (CVLT) or Rey Auditory Verbal Learning Test (RAVLT) yield accuracies of over 80% when predicting conversion from mild cognitive impairment (MCI) to AD (Eckerström et al., 2013; Rabin et al., 2009). However, predicting future progression or diagnosis from NC (or even MCI and dementia) is difficult due to clinician subjectivity and individual patient variability. The implementation of machine learning techniques to analyze AD-related biomarkers may help improve early detection models and increase classification accuracy. Furthermore, the use of imaging techniques as opposed to cognitive tests may improve accuracies because they are not influenced by clinician subjectivity and less influenced by patient day-to-day variability.

Using MRI, researchers can measure neurodegeneration by analyzing volumetric changes in older adults’ brains. When studying changes due to aging, MCI, and AD, the most commonly studied area is the hippocampus because this region experiences atrophy early in the disease course (Fjell et al., 2014). The hippocampus has also been identified as one of the most useful biomarkers when examining progression from MCI to AD (Risacher et al., 2009). Another method of measuring hippocampal differences between groups is hippocampal grading, measured by the Scoring by Nonlocal Image Patch Estimator (SNIPE) (Coupe et al., 2015; Coupé, Eskildsen, Manjón, Fonov, & Collins, 2012; Coupé, Eskildsen, Manjón, Fonov, Pruessner, et al., 2012). This method has proven to be associated with cognitive changes in cognitive healthy older adults and people with early MCI (Morrison et al., 2022). Furthermore, this method has proven to classify between cognitively healthy older adults and people with AD with an accuracy of 93% when using both the hippocampus and entorhinal cortex (Coupé, Eskildsen, Manjón, Fonov, & Collins, 2012). However, these SNIPE results come from limit samples and need to be further examined to determine their usefulness in classifying different disease cohorts.

To better understand the potential of SNIPE to correctly classify aging and cognitive impairment groups more research is needed on 1) larger samples and in people with MCI, 2) people who are amyloid positive and are thus on the AD trajectory, and 3) comparing SNIPE to established methods such as Freesurfer. A larger sample is needed in these machine learning studies to reduce the chance of overfitting and to improve generalizability to other samples. Coupé and colleagues’ papers examining SNIPE used the Alzheimer’s Disease Neuroimaging Initiative diagnosis label ‘AD’ to differentiate groups (2012, 2015). The problem with the ADNI AD diagnosis is that it is based solely on clinical scores and does not include amyloid positivity. Based on the National Institute on Aging – Alzheimer’s Association biomarker AD profiles, an older adult with abnormal amyloid levels (amyloid positive) is placed in the Alzheimer’s continuum, whereas someone without amyloid positivity is either experiencing non-AD pathologic change or has normal-level AD biomarkers (Jack, 2018). Thus, in order to correctly classify people as AD or as NC on the disease continuum it is important to examine only those who are amyloid positive. Specifically examining people on the AD continuum is also necessary because they are often selected for clinical trials and their accurate classification has the potential to further improve clinical trial patient selection. Finally, while the current research on SNIPE has offered promising results, the findings have yet to be compared to the traditional methods used to measure hippocampal volume (i.e., Freesurfer).

The goal of this study was to determine whether classification accuracy is higher for SNIPE hippocampal grading, compared to SNIPE hippocampal volume, and Freesurfer volume. A recent review has also shown that few studies compute classifications between MCI vs AD, with most studies focusing on classifying NC from AD (Tanveer et al., 2020). While the former is a bit late for early intervention, the latter is not really of clinical interest. Therefore, we designed this study to examine classification accuracy between healthy older adults, people early mild cognitive impairment (eMCI) and late MCI (lMCI), and people with AD. Several commonly used classifiers were applied to determine which technique (i.e., SNIPE Grading, SNIPE volume or FreeSurfer volume) is best at classifying participants into their correct diagnostic cohort. Including multiple techniques improves the generalizability of our results and comparison to past research.

## 2 Methods

### 2.1 Alzheimer’s Disease Neuroimaging Initiative

Data used in the preparation of this article were obtained from the Alzheimer’s Disease Neuroimaging Initiative (ADNI) database (adni.loni.usc.edu). The ADNI was launched in 2003 as a public-private partnership, led by Principal Investigator Michael W. Weiner, MD. The primary goal of ADNI has been to test whether serial magnetic resonance imaging (MRI), positron emission tomography (PET), other biological markers, and clinical and neuropsychological assessment can be combined to measure the progression of mild cognitive impairment (MCI) and early Alzheimer’s disease (AD). Participants were selected from ADNI-1, ADNI-2, and the ADNI-GO cohorts. The study received ethical approval from the review boards of all participating institutions. Written informed consent was obtained from participants or their study partner.

All ADNI participants were imaged using a 3T scanner with T1-weighted imaging parameters (see http://adni.loni.usc.edu/methods/mri-tool/mri-analysis/ for the detailed MRI acquisition protocol). Baseline scans were downloaded from the ADNI public website.

### 2.2 Participants from ADNI cohort

Participant inclusion and exclusion criteria are available at www.adni-info.org. All participants were between the ages of 55 and 90 at the time of recruitment, exhibiting no evidence of depression. Healthy normal controls had no evidence of memory decline, as measured by the Wechsler Memory Scale and no evidence of impaired global cognition as measured by the Mini Mental Status Examination (MMSE) or Clinical Dementia Rating (CDR). Both eMCI and lMCI had scores between 24 and 30 on the MMSE, 0.5 on the CDR, and abnormal scores on the Wechsler Memory Scale. AD was defined as participants who had abnormal memory function on the Wechsler Memory Scale, an MMSE score between 20 and 26, a CDR of 0.5 or 1.0 and probable AD according to the NINCDS/ADRDA criteria.

Figure 1 summarizes the methodology used to select participants from the ADNI studies. A total of 1666 participants were selected from ADNI-1 (n= 799), ADNI-2 (n=776), ADNI-GO (n=91) who had baseline MRI scans that passed quality control, for which hippocampal grading and volume could be extracted (513 NC, 269 eMCI, 556 lMCI, 328 AD).

**Figure 1:**
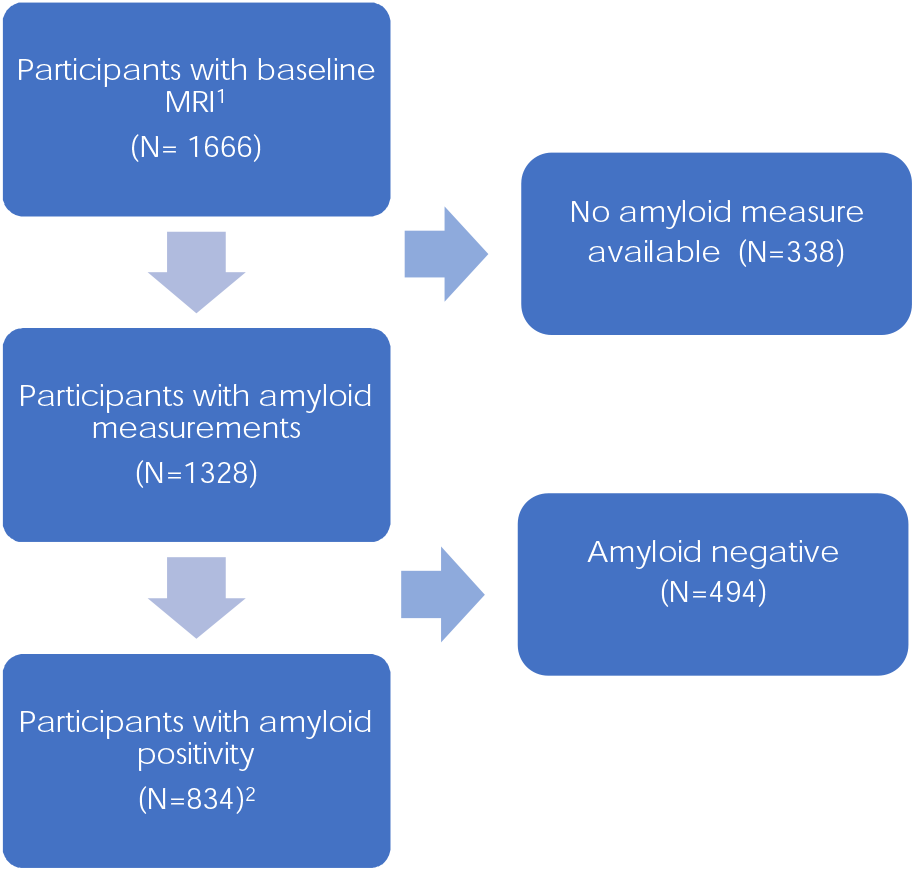
Flowchart summarizing the participant inclusion and exclusion criteria based on amyloid positivity. *Notes:* ^1^ = the participants included in the first analysis including the entire sample. ^2^ = amyloid positive participants included in the second analysis focusing on those in the AD trajectory

Amyloid status was derived from PET or CSF measures. ADNI PET data was acquired on multiple PET instruments with different acquisition sequences following various platform-specific acquisition protocols. All PET data underwent quality control and standard image pre-processing correct steps to improve data uniformity across collection sites. More detail can be downloaded from the ADNI procedures manual. The AV-45 PET scans were collected approximately 50 minutes post injection (Landau and Jagust 2015). The PiB-PET scans were collected after 50-70 minutes post injection of approximately 15 mCi (Jagust et al., 2010). To obtain cerebrospinal fluid (CSF) samples, lumbar punctions were performed as described in the ADNI procedures manual. CSF Aß42 were measured using the multiplex xMAP Luminex platform (Luminex Corp, Austin, TX, USA) with the INNO-BIA AlzBio3 kit (Innogenetics) (Olsson et al., 2005; Shaw et al., 2009).

The current definition of dementia in ADNI is solely based on clinical assessments and does not include amyloid positivity. For this reason, we also wanted to repeat the same analysis in a subset of amyloid positive participants to ensure we are testing on people who are experiencing Alzheimer’s related pathological changes. To determine amyloid positivity, both CSF and PET values were used as not all participants had both measurements available. Participants were identified as amyloid positive if they had any of the following at baseline: 1) a standardized update value ratio (SUVR) of > 1.11 on AV45 PET (Landau et al., 2013), 2) a SUVR of >1.2 using Pittsburgh compound-B PET (Villeneuve et al., 2015), or 3) a cerebrospinal fluid Aß1-42 ≤ 980 pg/ml as per ADNI recommendations. Of the 1666 participants selected, 1328 had baseline amyloid levels available to determine amyloid positivity (427 NC, 263 eMCI, 400 lMCI, 238 AD) and of those 834 were amyloid positive (179 NC, 148 eMCI, 298 lMCI, 209 AD).

### 2.3 Participants from AIBL cohort

Participant inclusion and exclusion criteria for the Australian Imaging, Biomarker & Lifestyle Flagship Study of Ageing (AIBL) have been previously full described (Ellis et al., 2009). Briefly, healthy controls were 60+ and in good general health with no evidence of cognitive impairment. Those with MCI had to score <28/30 on the MMSE, and have abnormal scores on the Wechsler Memory Scale, and a CDR score of 0.5 or greater. AD patients were characterized by the NINCDS-ADRDA criteria (McKhann et al., 1984). There were 858 participants, of these participants only 581 participants had baseline MRIs that passed quality control for which hippocampal grading and volume could be extracted were included (413 NC, 90 MCI, 78 AD).

### 2.4 Structural MRI processing

Raw T1w scans for each participant were pre-processed through our standard pipeline including noise reduction (Coupe et al., 2008), intensity inhomogeneity correction (Sled, Zijdenbos, & Evans, 1998), and intensity normalization into the range 0-100. The pre-processed images were then linearly (9 parameters: 3 translation, 3 rotation, and 3 scaling) (Dadar et al., 2018) registered to the MNI-ICBM152-2009c average (Fonov et al., 2011). The quality of the linear registrations was visually verified by an experienced rater (author M.D.), blinded to diagnostic group. Only seven datasets did not pass this quality control step and were discarded.

### 2.5 SNIPE grading and volume

Scoring by Nonlocal Image Patch Estimator (SNIPE) was used to segment the hippocampus and measure the extent of AD-related change within the hippocampus using the linearly registered preprocessed T1-weighted images (Coupé, Eskildsen, Manjón, Fonov, & Collins, 2012; Coupé, Eskildsen, Manjón, Fonov, Pruessner, et al., 2012). The SNIPE procedure used has been previously described in detail (Dadar et al., 2020). In this method (SNIPE), volumes are calculated by counting voxels in a pseudo-Talairach stereotaxic space, thus correcting for subject’s head size. The quality of the SNIPE segmentations were visually verified (author N.S.), blinded to diagnostic group. Only 11 datasets did not pass this quality control step and were discarded from the analyses.

### 2.6 FreeSurfer volumes

Freesurfer volumes were used to complete the classification analysis with the ADNI data. The volumes were provided by ADNI, completed using the standard protocols developed and implemented by The University of California, San Francisco (UCSF). For a more detailed explanation of the pre-processing and quality control guidelines, please see the full UCSF FreeSurfer Overview and QC Guide.

### 2.7 Data availability statement

The data used for this analysis (both AIBL and ADNI data) are available on request from the ADNI database (ida.loni.usc.edu).

### 2.8 Classification Analysis

To assess the prediction power of each measure, a classification scheme with 10-fold cross validation was used. Specifically, SNIPE grading, SNIPE volume, or Freesurfer volume scores for the left and right HC were summed and used as features along with participant age and sex. To ensure that the potential differences in the distribution of the random splits in the cross-validation folds do not impact the results, the same splits were consistently used for assessment of the performance of the three features evaluated with a support vector machine (SVM) classifier. To facilitate comparison with other studies in the literature and to ensure that the results are not classifier dependent, six other classifiers were examined: decision tree, k-nearest neighbors, error-correcting output codes, binary Gaussian kernel, binary linear, and random forests. The default parameters were used for each classifier. All analyses were performed using MATLAB version 9.7.

Independent validation of the classification was completed using NC, MCI, and AD participants from the AIBL cohort. While ADNI classifies MCI participants into either eMCI or lMCI, AIBL only uses MCI. For that reason classification models for NC:MCI and MCI:AD were created using only the ADNI training set: 1) with both eMCI and lMCI participants, 2) with only the eMCI participants, and 3) with only the lMCI participants. Importantly, AIBL data was used only for independent validation of these three models. No AIBL data was used in the creation of these models, nor for any parameter adjustment.

### 2.9 Statistical Analysis

Demographic information between groups was compared using independent sample t-tests and corrected for multiple comparisons using Bonferroni correction.

## 3 Results

### 3.1 Demographic and clinical results in ADNI

Table 1 provides the demographic characteristics for all participants separated by group for both the full sample and amyloid positive sub-group.

**Table 1:** Demographic information for cognitively normal, early and late MCI, and AD participants.

| *Full Sample* | **NC**<br>**n=513** | **eMCI**<br>**n= 269** | **IMCI**<br>**n= 556** | **AD**<br>**n= 328** |
| --- | --- | --- | --- | --- |
| Age | 74.36 ± 5.79 | 70.82 ± 7.38 | 73.99 ± 7.53 | 75.00 ± 7.78 |
| Education | 16.35 ± 2.71 | 15.93 ± 2.65 | 15.90 ± 2.92 | 15.16 ± 3.01 |
| Female Sex | 265 (52%) | 120 (45%) | 215 (39%) | 147 (45%) |
| ADAS-13 | 9.30 ± 4.54 | 12.59 ± 5.54 | 18.70 ± 6.53 | 29.95 ± 7.90 |
| CDR-SB | 0.05 ± 0.19 | 1.29 ± 0.75 | 1.65 ± 0.92 | 4.45 ± 1.62 |
| *Amyloid Positive* | **NC**<br>**n=179** | **eMCI**<br>**n=148** | **IMCI**<br>**n=298** | **AD**<br>**n=209** |
| Age | 74.81 ± 5.68 | 72.29 ± 7.11* | 73.51 ± 7.14 | 74.16 ± 8.01 |
| Education | 16.22 ± 2.67 | 15.83 ± 2.74 | 16.00 ± 2.87 | 15.44 ± 2.79 |
| Female Sex | 86 (62%) | 62 (42%) | 121 (41%) | 93 (44%) |
| ADAS-13 | 9.64 ± 4.62 | 13.70 ± 5.46 | 19.77 ± 6.65 | 30.74 ± 8.21 |
| CDR-SB | 0.06 ± 0.27 | 1.40 ± 0.84 | 1.70 ± 0.91 | 4.53 ± 1.60 |

*Notes.* Values are expressed as mean ± standard deviant, or number (percentage %). Female Sex is represented as total number of sample and percentage of sample. NC = cognitively normal controls. eMCI = early mild cognitive impairment. IMCI = late mild cognitive impairment. AD = Alzheimer's disease. ADAS-13 = Alzheimer's Disease Assessment Scale–Cognitive Subscale. CDRSB = Clinical Dementia Rating Scale – Sum of Boxes. \* eMCI were younger than NC.

Several demographic and clinical factors differed between sub-groups in the full sample after correction for multiple comparison. With respect to age differences, eMCI were significantly younger than NC (*t*=6.84, *p*<.001), people with lMCI (*t*=5.74, *p*<.001), and people with AD (*t*=6.71, *p*<.001). AD had significantly lower education than NC (*t*=5.84, *p*<.001), people with eMCI (*t*=3.54, *p*<.001), and lMCI (*t*=3.59, *p*<.001). All groups significantly differed in ADAS-13 scores, with scores progressively increasing with each stage of decline (NC:eMCI, *t*= -8.31, *p*<.001; eMCI:lMCI, *t*= -13.89, *p*<.001; and lMCI:AD, *t*= -21.51, *p*<.001). Similarly, all groups significantly differed in CDR-SB scores, with scores progressively increasing with each stage of decline (NC:eMCI, *t*=-26.44, *p*<.001; eMCI:lMCI, *t*=-6.01, *p<*.001; and lMCI:AD, *t*=-28.51, *p*<.001).

Demographic differences were also observed in the amyloid positive sub-analysis. After correction for multiple comparisons, eMCI were younger than only NC (*t*=3.49, *p*<.001). No group differences were observed in education after correction for multiple comparisons. All groups significantly differed in ADAS-13 scores, with scores progressively increasing with each stage of decline (NC:eMCI, *t*= -7.12, *p*<.001; eMCI:lMCI, *t*= -10.16, *p*<.001; and lMCI:AD, *t*= - 15.83, *p*<.001). Similarly, all groups significantly differed in CDR-SB scores, with scores progressively increasing with each stage of decline (NC:eMCI, *t*=-18.34, *p*<.001; eMCI:lMCI, *t*=-3.49, *p<*.001; and lMCI:AD, *t*=-24.07, *p*<.001).

### 3.2 Classification results in ADNI

Table 2 shows the participant classification accuracy, sensitivity, and specificity scores for SNIPE grading, SNIPE volume, and Freesurfer volume for each analysis using SVM. Accuracy, sensitivity, and specificity obtained using the other classifiers are available in Supplementary Table S.1.

**Table 2:** Percent accuracy, sensitivity, and specificity (± standard deviation, evaluated over 10 folds) for classifying cognitively normal, early and late MCI, and AD participants.

| **SVM** | NC: AD |  |  | NC: eMCI |  |  | eMCI: lMCI |  |  | lMCI: AD |  |  |
| --- | --- | --- | --- | --- | --- | --- | --- | --- | --- | --- | --- | --- |
|  | Grading | Volume | Freesurfer | Grading | Volume | Freesurfer | Grading | Volume | Freesurfer | Grading | Volume | Freesurfer |
| Accuracy | 89 ± 4 | 80 ± 4 | 65 ± 12 | 70 ± 4 | 68 ± 3 | 62 ± 6 | 67 ± 5 | 67 ± 5 | 53 ± 14 | 68 ± 6 | 63 ± 5 | 60 ± 7 |
| Sensitivity | 82 ± 6 | 66 ± 6 | 30 ± 3 | 23 ± 8 | 15 ± 4 | 37 ± 2 | 89 ± 8 | 100 ± 0 | 49 ± 3 | 45 ± 6 | 0 ± 0 | 24 ± 3 |
| Specificity | 94 ± 5 | 89 ± 8 | 87 ± 8 | 94 ± 4 | 96 ± 2 | 76 ± 17 | 23 ± 2 | 0 ± 0 | 60 ± 2 | 82 ± 5 | 100 ± 0 | 82 ± 2 |
| **Amyloid Positive** | Grading | Volume | Freesurfer | Grading | Volume | Freesurfer | Grading | Volume | Freesurfer | Grading | Volume | Freesurfer |
| Accuracy | 87 ± 2 | 76 ± 5 | 46 ± 11 | 63 ± 6 | 59 ± 5 | 56 ± 10 | 67 ± 5 | 67 ± 4 | 51 ± 14 | 67 ± 4 | 58 ± 4 | 57 ± 4 |
| Sensitivity | 85 ± 5 | 77 ± 5 | 26 ± 3 | 52 ± 6 | 48 ± 2 | 32 ± 2 | 88 ± 8 | 100 ± 0 | 44 ± 3 | 57 ± 9 | 0 ± 0 | 23 ± 3 |
| Specificity | 89 ± 3 | 75 ± 5 | 68 ± 9 | 72 ± 4 | 69 ± 1 | 77 ± 2 | 27 ± 2 | 0 ± 0 | 65 ± 3 | 74 ± 6 | 100 ± 0 | 81 ± 15 |

Notes: SVM = support vector machines, NC = normal controls, AD= Alzheimer's disease, eMCI = early mild cognitive impairment. lMCI = late mild cognitive impairment, Grading = Scoring by Nonlocal Image Patch Estimator Hippocampal Grading, Volume = the Scoring by Nonlocal Image Patch Estimator Hippocampal Volume, Freesurfer = Hippocampal volume measured with Freesurfer.

The highest classification accuracy was obtained using SNIPE grading, followed by SNIPE volume, then Freesurfer. This order was obtained for all group classifications in both the full sample and amyloid positive sample. When examining NC vs. AD classification, SNIPE grading provided an accuracy of 89% compared to 80% for SNIPE volume and 65% obtained using Freesurfer. When examining NC vs. AD in the amyloid positive sample SNIPE grading accuracy dropped by only 2% to 87%. On the other hand, SNIPE volume accuracy dropped by 4% to 76% and Freesurfer classification accuracy dropped to 46%, a difference of 19%. In the NC vs. eMCI classification, SNIPE grading obtained the highest with 70% accuracy compared to SNIPE volume with 68% and Freesurfer volume with only 62%. When comparing NC vs. eMCI in the amyloid positive sample SNIPE grading obtained the highest accuracy compared to SNIPE volume and Freesurfer volume (63% vs 59% and 56%, respectively).

In the eMCI vs. lMCI classification, SNIPE grading and SNIPE volume obtained the highest with 67% accuracy compared to Freesurfer volume with 53%. When comparing eMCI vs. lMCI in the amyloid positive sample SNIPE grading and volume accuracies did not change while Freesurfer accuracy dropped 2% to 51%. In the lMCI vs. AD classification, SNIPE grading obtained the highest with 68% accuracy compared to SNIPE volume at 63% and Freesurfer with 60%. When comparing lMCI vs. AD in the amyloid positive sample SNIPE grading once again provided the highest classification accuracy compared to SNIPE volume and Freesurfer volume (67% vs 58% and 57%, respectively).

It is interesting to note that in all experiments, the standard deviation of the Freesurfer volume classification accuracy is much larger that SNIPE grading or SNIPE volumes except for lMCI:AD.

### 3.3 Classification results in the AIBL cohort (external validation)

In the first set of classification results we observed that SNIPE grading and volume were both more accurate than using hippocampal volumes obtained using Freesurfer at classifying groups. For that reason, in this validation procedure only SNIPE grading and volume were compared because both these techniques outperformed hippocampal volumes obtained using Freesurfer. Furthermore, given the similar prediction accuracies between the different classifiers, this external validation was completed using only the SVM classifier. Table 3 shows the participant classification accuracy, sensitivity, and specificity scores for SNIPE grading and SNIPE volume for each analysis using SVM.

**Table 3:** Percent accuracy, sensitivity, and specificity for classifying NC, MCI and AD participants in the AIBL cohort.

| **SVM** | NC:AD |  | NC:MCI<sup>1</sup> |  | NC:MCI<sup>2</sup> |  | NC:MCI<sup>3</sup> |  | MCI:AD<sup>1</sup> |  | MCI:AD<sup>2</sup> |  | MCI:AD<sup>3</sup> |  |
| --- | --- | --- | --- | --- | --- | --- | --- | --- | --- | --- | --- | --- | --- | --- |
|  | Grading | Volume | Grading | Volume | Grading | Volume | Grading | Volume | Grading | Volume | Grading | Volume | Grading | Volume |
| Accuracy | 90 | 80 | 55 | 41 | 79 | 82 | 74 | 64 | 66 | 54 | 65 | 59 | 66 | 54 |
| Sensitivity | 77 | 59 | 67 | 79 | 27 | 0 | 56 | 59 | 29 | 0 | 76 | 76 | 40 | 0 |
| Specificity | 93 | 88 | 53 | 32 | 90 | 100 | 78 | 65 | 93 | 100 | 57 | 46 | 89 | 100 |

Notes: AIBL = Australian Imaging, Biomarker & Lifestyle Flagship Study of Ageing. SVM = support vector machines, NC = normal controls, MCI = mild cognitive impairment, AD= Alzheimer's disease. Grading = Scoring by Nonlocal Image Patch Estimator Hippocampal Grading, Volume = the Scoring by Nonlocal Image Patch Estimator Hippocampal Volume. <sup>1</sup> = Training sample from ADNI used both eMCI and lMCI. <sup>2</sup> = Training sample from ADNI used only eMCI. <sup>3</sup> = Training sample from ADNI used only lMCI.

The highest classification accuracy was obtained using SNIPE grading over SNIPE volume for almost all analyses. When examining NC vs. AD classification, SNIPE grading provided an accuracy of 90% compared to 80% for SNIPE volume. In the NC:MCI prediction, accuracy for SNIPE grading was higher than SNIPE volume 55% vs 41% (trained with eMCI and lMCI) and 74% vs 64% (trained with lMCI only), but higher for SNIPE volume over SNIPE grading 82% vs 79% (trained with eMCI only). In the MCI:AD prediction, accuracy for SNIPE grading was higher than SNIPE volume in all three training cases, 66% vs 54% (trained with eMCI and lMCI), 65% vs 59% (trained with eMCI only), 66 vs 54% (trained with lMCI only).

## 4 Discussion

In recent years, numerous studies have implemented machine learning techniques on imaging data with the goal of accurately classifying and predicting people with dementia and more specifically AD (see Tanveer et al., 2020 for review). Many of these studies have been completed on small samples, do not attempt to classify people with MCI (from NC or AD), and relatively few have implemented new techniques aimed at measuring changes in the hippocampus. The current study addressed these limitations by comparing classification accuracy on a large sample of NC, and people with eMCI, lMCI, and AD (and with MCI and AD in AIBL), using a relatively new method to detect hippocampal changes. The findings observed here show that SNIPE grading has higher classification accuracy than both SNIPE volume and Freesurfer volume when classifying: 1) NC:AD, 2) NC:eMCI, 3) eMCI:lMCI, and 4) lMCI:AD in both the full sample and the amyloid positive sub-sample of ADNI and when classifying 1) NC:AD, 2) NC:MCI and 3) MCI:AD in the AIBL cohort..

These findings compliment those previously completed on SNIPE grading and volume (Coupe et al., 2012a,b, 2015). They found that SNIPE grading could classify between NC and AD with 93% accuracy using both hippocampal and entorhinal cortex grading (Coupe et al., 2012a). Coupe et al., (2012b) also observed that grading was more accurate than volume at classifying progressive MCI vs stable MCI. The current study also found high accuracy (89%) when classifying NC vs. AD using only hippocampal grading and observed the novel finding that when focusing on those in the AD trajectory (amyloid positive) the results also remained high (87%). This finding of high accuracy in the amyloid positive group is important when attempting to identify those in the pre-clinical stages of AD. High accuracy at identifying those in the pre-clinical stage of AD is important for clinical trials and further shows the importance of these results. Future work should determine the predictability of SNIPE at determining which amyloid positive NC will convert to pathological AD.

Compared to Freesurfer, SNIPE measures provided higher classification accuracy for all group classifications. The comparison of SNIPE to Freesurfer is essential in determining the usefulness of SNIPE because Freesurfer is a common method used to examine volume changes in AD. A quick MEDLINE search shows almost 600 papers with the keywords of Freesurfer and AD. Furthermore, Freesurfer volumetric measures are provided for the hippocampus in ADNI. While Freesurfer is somewhat accurate at classifying NC vs. AD (65%), Grading was much more accurate (89%), with a 24% improvement in classification accuracy. Similarly, differentiating between NC vs. AD in amyloid positive group the classification was much more robust with Grading compared to Freesurfer (87% vs. 52%), with grading offering 35% higher accuracy. These findings show that Grading is more sensitive than Freesurfer to hippocampal differences that occur in AD compared to NC. It should be noted that while we use the term AD to refer to both the full-sample based on clinical diagnosis and the amyloid positive sub-sample, the full sample represents AD and other dementias. Thus, these findings suggest that not only is grading accurate at detecting to hippocampal changes due to dementia but is also highly accurate at classifying between NC and AD who are on the AD path. An important benefit of using the SNIPE grading scores proposed here is the robustness of the method. In the 1666 MRI volumes processed, only 7 failed stereotaxic registration and 11 failed SNIPE segmentation. Robustness is important in clinical trials since losing data to pipeline failures results in reduced power to detect group differences. Another advantage of the SNIPE method is the low standard deviation compared to Freesurfer in all the classification analyses.

Similar classification accuracies were also obtained when validating our results in the independent AIBL cohort. We obtained a high accuracy of 90% when classifying NC vs. AD in the AIBL cohort. Accuracies similar to those observed using the ADNI dataset were also observed in the AIBL NC vs. MCI and MCI vs. AD classification. These findings suggest that the classification model is not only accurate in the ADNI dataset but is generalizable to and replicable in other datasets. In a recent meta-analysis, authors found that almost 30% of articles reviewed use the test set in the training process, thus *double dipping* during their evaluation (Ansart et al., 2021). This finding further emphasizes the importance of the current study using cross-validation in the original dataset as well as using an independent cohort for validation.

The results found here are better or equivalent to past machine learning techniques that attempt to classify different disease cohorts from each other and NCs (see Taveer et al., 2020). The majority of the studies examined in the above-mentioned review focused on only classifying NC vs. AD, with less than 25% of the 60 studies using SVM to classify between MCI and AD and just over 30% classifying between NC and MCI (Taveer et al., 2020). In order to target early diagnosis of AD, researchers must be able to correctly classify between MCI and AD. In our sample, we were able to differentiate between eMCI vs. lMCI and lMCI vs. AD with almost 70% accuracy. Although there is some research that has shown similar success the novelty and advantage of the current results is that we employed a larger sample to improve generalizability, examined those on the AD trajectory (by studying amyloid positive groups), and classified people with eMCI vs. lMCI. Furthermore, we also validated these results in an independent cohort of NCs, MCI, and AD. These findings show promise for the use of SNIPE grading as a powerful MRI-based feature that could be used in conjunction with other data to improve classification of patients into the correct disease cohort. Future research should examine whether SNIPE grading is useful at detecting which amyloid positive subjects will cognitively decline and develop AD. Early detection of AD will not only improve a clinicians’ ability to provide effective care to patients but also potentially improve selection of patients for clinical trials.

## 5 Conclusion

The current paper observed that SNIPE grading scores provided higher classification accuracy than both SNIPE volume and Freesurfer volumes. Importantly, this classification accuracy remained similar in the independent validation analysis using the AIBL cohort. These findings suggest that HC grading offers promise as a method to accurate classify those with and without AD. Future work should examine whether HC grading is predictive of future conversion from NC to MCI and dementia.

## Data Availability

All data produced in the present study are available upon written request to adni.loni.usc.edu.

https://adni.loni.usc.edu/

## Acknowledgments

Data collection and sharing for this project was funded by the Alzheimer’s Disease Neuroimaging Initiative (ADNI) (National Institutes of Health Grant U01 AG024904) and DOD ADNI (Department of Defense award number W81XWH-12-2-0012). ADNI is funded by the National Institute on Aging, the National Institute of Biomedical Imaging and Bioengineering, and through generous contributions from the following: AbbVie, Alzheimer’s Association; Alzheimer’s Drug Discovery Foundation; Araclon Biotech; BioClinica, Inc.; Biogen; Bristol-Myers Squibb Company; CereSpir, Inc.; Cogstate; Eisai Inc.; Elan Pharmaceuticals, Inc.; Eli Lilly and Company; EuroImmun; F. Hoffmann-La Roche Ltd and its affiliated company Genentech, Inc.; Fujirebio; GE Healthcare; IXICO Ltd.; Janssen Alzheimer Immunotherapy Research & Development, LLC.; Johnson & Johnson Pharmaceutical Research & Development LLC.; Lumosity; Lundbeck; Merck & Co., Inc.; Meso Scale Diagnostics, LLC.; NeuroRx Research; Neurotrack Technologies; Novartis Pharmaceuticals Corporation; Pfizer Inc.; Piramal Imaging; Servier; Takeda Pharmaceutical Company; and Transition Therapeutics. The Canadian Institutes of Health Research is providing funds to support ADNI clinical sites in Canada. Private sector contributions are facilitated by the Foundation for the National Institutes of Health (www.fnih.org). The grantee organization is the Northern California Institute for Research and Education, and the study is coordinated by the Alzheimer’s Therapeutic Research Institute at the University of Southern California. ADNI data are disseminated by the Laboratory for Neuro Imaging at the University of Southern California.

