## Supplemental Table1 for "Hippocampal grading provides higher Alzheimer’s Disease prediction accuracy than hippocampal volume"

Supplementary Table 1: Percent accuracy, sensitivity, and specificity for classifying cognitively normal, early and late MCI, and AD participants.

| **Decision Tree** | NC:AD | | |  | | NC:eMCI | | | | | | |  | | eMCI:lMCI | | | |  | | lMCI:AD | | | | | |
| --- | --- | --- | --- | --- | --- | --- | --- | --- | --- | --- | --- | --- | --- | --- | --- | --- | --- | --- | --- | --- | --- | --- | --- | --- | --- | --- |
| *Full sample* | Grading | Volume | Freesurfer | |  | | Grading | | Volume | | Freesurfer | | |  | | Grading | Volume | Freesurfer | |  | | Grading | Volume | | Freesurfer | |
| Accuracy | 87 | 74 | 77 | |  | | | 66 | | 65 | | 63 | |  | | 66 | 65 | 64 | |  | | 58 | | 56 | | 59 |
| Sensitivity | 83 | 66 | 60 | |  | | | 48 | | 42 | | 46 | |  | | 76 | 77 | 76 | |  | | 44 | | 35 | | 31 |
| Specificity | 89 | 79 | 88 | |  | | | 75 | | 78 | | 72 | |  | | 46 | 41 | 40 | |  | | 67 | | 69 | | 75 |
| *Amyloid Positive* | Grading | Volume | Freesurfer | |  | | | Grading | | Volume | | Freesurfer | |  | | Grading | Volume | Freesurfer | |  | | Grading | | Volume | | Freesurfer |
| Accuracy | 83 | 67 | 76 | |  | | | 65 | | 59 | | 61 | |  | | 63 | 65 | 64 | |  | | 53 | | 51 | | 56 |
| Sensitivity | 84 | 70 | 85 | |  | | | 60 | | 57 | | 55 | |  | | 73 | 76 | 75 | |  | | 48 | | 35 | | 37 |
| Specificity | 83 | 63 | 66 | |  | | | 69 | | 61 | | 65 | |  | | 43 | 44 | 42 | |  | | 57 | | 62 | | 69 |
| **Error-correcting output codes** | NC:AD | | |  | | NC:eMCI | | | | | | |  | | eMCI:lMCI | | | |  | | lMCI:AD | | | | | |
| *Full sample* | Grading | Volume | Freesurfer | |  | | Grading | | Volume | | Freesurfer | | |  | | Grading | Volume | Freesurfer | |  | | Grading | Volume | | Freesurfer | |
| Accuracy | 89 | 80 | 71 | |  | | | 70 | | 68 | | 62 | |  | | 67 | 67 | 32 | |  | | 68 | | 63 | | 61 |
| Sensitivity | 82 | 66 | 46 | |  | | | 23 | | 15 | | 37 | |  | | 88 | 99 | 15 | |  | | 45 | | 0 | | 11 |
| Specificity | 94 | 89 | 87 | |  | | | 94 | | 97 | | 75 | |  | | 25 | 1.5 | 70 | |  | | 82 | | 100 | | 90 |
| *Amyloid Positive* | Grading | Volume | Freesurfer | |  | | | Grading | | Volume | | Freesurfer | |  | | Grading | Volume | Freesurfer | |  | | Grading | | Volume | | Freesurfer |
| Accuracy | 86 | 75 | 51 | |  | | | 63 | | 59 | | 53 | |  | | 67 | 67 | 39 | |  | | 67 | | 59 | | 56 |
| Sensitivity | 83 | 75 | 25 | |  | | | 52 | | 48 | | 33 | |  | | 88 | 100 | 25 | |  | | 57 | | 0 | | 27 |
| Specificity | 89 | 89 | 81 | |  | | | 72 | | 69 | | 69 | |  | | 26 | 0 | 68 | |  | | 74 | | 100 | | 76 |

| **Gaussian Kernel** | NC:AD | | |  | | NC:eMCI | | | | | | |  | | eMCI:lMCI | | | |  | | lMCI:AD | | | | | |
| --- | --- | --- | --- | --- | --- | --- | --- | --- | --- | --- | --- | --- | --- | --- | --- | --- | --- | --- | --- | --- | --- | --- | --- | --- | --- | --- |
| *Full sample* | Grading | Volume | Freesurfer | |  | | Grading | | Volume | | Freesurfer | | |  | | Grading | Volume | Freesurfer | |  | | Grading | Volume | | Freesurfer | |
| Accuracy | 81 | 72 | 59 | |  | | | 71 | | 68 | | 63 | |  | | 73 | 70 | 66 | |  | | 63 | | 62 | | 59 |
| Sensitivity | 68 | 49 | 6.7 | |  | | | 32 | | 25 | | 6.3 | |  | | 93 | 91 | 95 | |  | | 14 | | 12 | | 10 |
| Specificity | 90 | 86 | 92 | |  | | | 91 | | 91 | | 93 | |  | | 31 | 25 | 5.5 | |  | | 92 | | 92 | | 88 |
| *Amyloid Positive* | Grading | Volume | Freesurfer | |  | | | Grading | | Volume | | Freesurfer | |  | | Grading | Volume | Freesurfer | |  | | Grading | | Volume | | Freesurfer |
| Accuracy | 82 | 70 | 53 | |  | | | 66 | | 63 | | 50 | |  | | 66 | 66 | 63 | |  | | 56 | | 54 | | 51 |
| Sensitivity | 84 | 74 | 66 | |  | | | 59 | | 55 | | 35 | |  | | 88 | 90 | 87 | |  | | 25 | | 23 | | 23 |
| Specificity | 79 | 65 | 39 | |  | | | 72 | | 69 | | 62 | |  | | 22 | 18 | 15 | |  | | 77 | | 76 | | 72 |
| **Linear** | NC:AD | | |  | | NC:eMCI | | | | | | |  | | eMCI:lMCI | | | |  | | lMCI:AD | | | | | |
| *Full sample* | Grading | Volume | Freesurfer | |  | | Grading | | Volume | | Freesurfer | | |  | | Grading | Volume | Freesurfer | |  | | Grading | Volume | | Freesurfer | |
| Accuracy | 88 | 79 | 73 | |  | | | 65 | | 66 | | 66 | |  | | 68 | 67 | 67 | |  | | 69 | | 63 | | 63 |
| Sensitivity | 79 | 64 | 49 | |  | | | 1 | | 1 | | 0 | |  | | 90 | 100 | 97 | |  | | 41 | | 0 | | 0 |
| Specificity | 93 | 88 | 89 | |  | | | 99 | | 99 | | 100 | |  | | 22 | 0 | 6 | |  | | 85 | | 100 | | 100 |
| **Amyloid Positive** | Grading | Volume | Freesurfer | |  | | | Grading | | Volume | | Freesurfer | |  | | Grading | Volume | Freesurfer | |  | | Grading | | Volume | | Freesurfer |
| Accuracy | 85 | 76 | 66 | |  | | | 61 | | 58 | | 53 | |  | | 68 | 67 | 66 | |  | | 65 | | 59 | | 59 |
| Sensitivity | 85 | 74 | 64 | |  | | | 52 | | 47 | | 0 | |  | | 85 | 100 | 95 | |  | | 49 | | 0 | | 0 |
| Specificity | 85 | 79 | 68 | |  | | | 68 | | 67 | | 98 | |  | | 33 | 0 | 1 | |  | | 76 | | 100 | | 100 |
| **Random Forest** | NC:AD | | |  | | NC:eMCI | | | | | | |  | | eMCI:lMCI | | | |  | | lMCI:AD | | | | | |
| *Full sample* | Grading | Volume | Freesurfer | |  | | Grading | | Volume | | Freesurfer | | |  | | Grading | Volume | Freesurfer | |  | | Grading | Volume | | Freesurfer | |
| Accuracy | 89 | 76 | 78 | |  | | | 66 | | 65 | | 66 | |  | | 69 | 65 | 67 | |  | | 58 | | 57 | | 62 |
| Sensitivity | 96 | 67 | 75 | |  | | | 43 | | 44 | | 42 | |  | | 80 | 78 | 80 | |  | | 42 | | 33 | | 37 |
| Specificity | 91 | 81 | 81 | |  | | | 77 | | 76 | | 78 | |  | | 44 | 38 | 41 | |  | | 68 | | 71 | | 77 |
| **Amyloid Positive** | Grading | Volume | Freesurfer | |  | | | Grading | | Volume | | Freesurfer | |  | | Grading | Volume | Freesurfer | |  | | Grading | | Volume | | Freesurfer |
| Accuracy | 84 | 73 | 75 | |  | | | 63 | | 59 | | 62 | |  | | 64 | 67 | 65 | |  | | 59 | | 52 | | 53 |
| Sensitivity | 87 | 71 | 80 | |  | | | 55 | | 54 | | 54 | |  | | 76 | 79 | 78 | |  | | 48 | | 39 | | 34 |
| Specificity | 81 | 69 | 69 | |  | | | 69 | | 63 | | 69 | |  | | 39 | 43 | 41 | |  | | 66 | | 61 | | 67 |

Notes:NC = normal controls, AD= Alzheimer’s disease, eMCI = early mild cognitive impairment. lMCI = late mild cognitive impairment, Grading = Scoring by Nonlocal Image Patch Estimator Hippocampal Grading, Volume = the Scoring by Nonlocal Image Patch Estimator Hippocampal Volume, Freesurfer = Hippocampal volume measured with Freesurfer.
